# Association between climate and new daily diagnoses of COVID-19

**DOI:** 10.1101/2020.11.12.20230888

**Authors:** Camilla Mattiuzzi, Brandon M. Henry, Fabian Sanchis-Gomar, Giuseppe Lippi

## Abstract

**Background:** Although evidence is accumulating that climate conditions may positively or negatively influence the scale of coronavirus disease 2019 (COVID-19) outbreaks, uncertainty remains concerning the real impact of climate factors on viral transmission. Methods. The number of new daily cases of COVID-19 diagnosed in Verona (Italy) was retrieved from the official website of Veneto Region, while information on daily weather parameters in the same area was downloaded from IlMeteo website, a renowned Italian technological company specialized in weather forecasts. The search period ranged between March 1 to November 11, 2020. The number of new daily COVID-19 cases and meteorological data in Verona were correlated using both univariate and multivariate analysis.

**Results:** The number of daily COVID-19 diagnoses in Verona was positively associated with the number of days in lockdown and humidity, and inversely correlated with mean, min and max temperature, mean wind speed and number of days with rainfall. Days of lockdown, mean air temperature, humidity, mean wind speed and number of days with rainfall remained significantly associated in multivariate analysis. The four weather parameters contributed to explaining 61% of variance in new daily COVID-19 diagnoses. Each 1% increase in air temperature, 1% decrease in humidity, 1 km/h increase in wind speed and day with rainfall were independently associated with 1.0%, 0.3%, 1.2% and 5.4% reduction in new COVID-19 daily diagnoses. A significant difference was observed in values of all-weather parameters recorded in Verona between days with <100 or ≥100 new daily COVID-19diagnoses.

**Conclusions:** Climate conditions may play an essential role in conditions of viral transmission, and influence the likelihood or course of local outbreaks. Preventive measures, testing policies and hospital preparedness should be reinforced during periods of higher meteorological risk and in local environments with adverse climate conditions.

## Introduction

It has now become clear that inter-human transmission of severe acute respiratory distress syndrome coronavirus 2 (SARS-CoV-2) is primarily through virus-containing droplets, emitted from nose or mouth, especially when infected persons exhale large volumes of air (e.g., coughing, sneezing, shouting, singing or exercising) (1). Transmission is also possible via aerosol-generating activities, especially with prolonged exposure to infected subjects within enclosed and poorly ventilated spaces (1). It should also be noted that droplets emitted from SARS-CoV-2 infected subjects and containing the virus can land on surfaces and objects in such a manner that the virus can remain viable and transmittable for hours, mostly via involuntarily or unconscious transport of the infected material to nose, mouth or eyes.

Based on these preferential means of viral transmission, a vast array of measures have been adopted to limit the spread of coronavirus disease 2019 (COVID-19). These encompass early diagnosis of potentially infective cases (2), hand hygiene, ambient sanitization, widespread use of facial masks, as well as social distancing or isolation (3, 4). Identifying additional determinants that may influence (i.e., either boost or limit) viral transmission within the community is essential, as this would enable to plan specific interventions against high-risk conditions. With respect to patient characteristics, the risk of being severely infected seems higher in people aged ≥40 years, in males, in blacks, in those overweight or co-morbidities (5). Specific activities and environmental aspects are also important factors in community transmission, including meeting, traveling or dining with infected individuals, especially without wearing face masks, being a family member of infected patients (6), and remaining for long periods in close proximity to SARS-CoV-2 infected subjects who are not wearing a face mask in crowded and poorly ventilated environments (7). A large recent study, which investigated the behavioral patterns of as many as 98 million Americans with respect to the risk of being infected by SARS-CoV-2, identified locations by which transmission risk was most likely to occur (8). The study identified fitness centers, café and snack bars, hotels and motels, religious organizations, physicians’ offices, grocery, merchandise, sporting goods and pet stores as common sites of transmission.

Recent evidence has also suggested that weather conditions may positively or negatively contribute to the scale of SARS-CoV-2 outbreaks (9, 10). However, since uncertainty remains concerning the real impact of some climate factors on the onset and/or spread of local clusters or outbreaks of SARS-CoV-2 (11), we designed a retrospective study in which we correlated daily weather parameters recorded in Verona (Italy) with the number of new official diagnoses of COVID-19 within the same geographical location, since the beginning of the local outbreak in March 2020.

## Materials and Methods

To specifically address the impact of weather on COVID-19 spread, we retrieved official data on the number of new daily cases of COVID-19 diagnosed in Verona (Italy) from the official website of the Veneto Region, whilst information on daily weather parameters in the same region was downloaded from the IlMeteo website (https://www.ilmeteo.it/portale/archivio-meteo/Verona), a renewed Italian technological company specialized in provision of services and communication on weather forecasts. The search period ranged between the date of the first COVID-19 case diagnosed in Verona (i.e., March 1, 2020) and November 11, 2020. The number of new daily COVID-19 cases and meteorological data in Verona were imported into a Microsoft Excel file (Microsoft, Redmond, WA, United States). The correlation between the number of new daily COVID-19 diagnoses, the days of lockdown in the area (i.e., between March 9 and May 18, 2020) and climate parameters were analyzed using univariate (Spearman’s correlation) and multivariate (multiple linear regression after logarithmic data transformation) analyses. In this latter case, the number of new COVID-19 daily cases was set as dependent variable, while climate data found to be significantly associated in univariate analysis, were set as independent variables. The difference in values of weather parameters recorded in Verona between days with <100 or ≥100 new daily diagnoses of COVID-19 in the same region was assessed with Mann Whitney U or chi-squared (χ^2^) tests, as appropriate. All continuous variables were reported as mean and standard deviation (SD). The statistical analysis was carried out using Analyse-it (Analyse-it Software Ltd, Leeds, UK). The study was conducted in accordance with the Declaration of Helsinki, under the terms of relevant local legislation. This retrospective analysis was based on searches of unrestricted, publicly available databases, and thereby no informed consent or Ethical Committee approvals were required.

## Results

The results of our analysis are shown in Table 1. A significant association was found in univariate analysis between the daily number of new COVID-19 diagnoses in Verona, days of lockdown and all the weather parameters investigated (i.e., mean temperature, min temperature, max temperature, humidity, mean wind speed, and number of days with rainfall). More specifically, the number of new daily COVID-19 diagnoses in Verona was found to be positively associated with the number of days in lockdown and humidity, while it was inversely correlated with mean, min and max temperature, mean wind speed and number of days with rainfall. In multivariate analysis, days of lockdown, mean air temperature, humidity, mean wind speed and number of days with rainfall remained significantly associated with new daily COVID- 19 cases (Table 1). Overall, these four weather parameters contributed to explaining 61% of the variance in the number of new daily COVID-19 cases. More specifically, each 1% increase in air temperature, 1% decrease in humidity, 1 km/h increase in wind speed and day with rainfall were independently associated with a 1.0%, 0.3%, 1.2% and 5.4% reduction in the number of new daily COVID-19 cases, respectively. Notably, a significant difference was also observed in values of all-weather parameters recorded in Verona between days with <100 or ≥100 new daily COVID-19 diagnoses, as shown in Table 2.

**Table 1.** Univariate and Multivariate correlation between new daily diagnoses of coronavirus disease 2019 (COVID-19) in Verona and the weather parameters recorded in the same area, between March 1 and November 11, 2020.

| Parameter | Univariate (r) | Multivariate ($\beta$ coefficient) |
| --- | --- | --- |
| **Lockdown** | 0.39 (95% CI, 0.28 to 0.49; $p<0.001$) | 0.42 ($p<0.001$) |
| **Mean temperature (°C)** | -0.64 (95% CI, -0.71 to -0.56; $p<0.001$) | -3.63 ($p=0.014$) |
| **Min temperature (°C)** | -0.65 (95% CI, -0.71 to -0.57; $p<0.001$) | -0.23 ($p=0.643$) |
| **Max temperature (°C)** | -0.58 (95% CI, -0.66 to -0.49; $p<0.001$) | -1.81 ($p=0.131$) |
| **Humidity (%)** | 0.15 (95% CI, 0.03 to 0.27; $p=0.015$) | 1.44 ($p=0.019$) |
| **Mean wind speed (km/h)** | -0.21 (95% CI, -0.33 to -0.09; $p=0.001$) | -0.86 ($p=0.006$) |
| **Days of rain** | -0.20 (95% CI, -0.31 to -0.08; $p=0.002$) | -0.30 ($p=0.002$) |

**Table 2.**
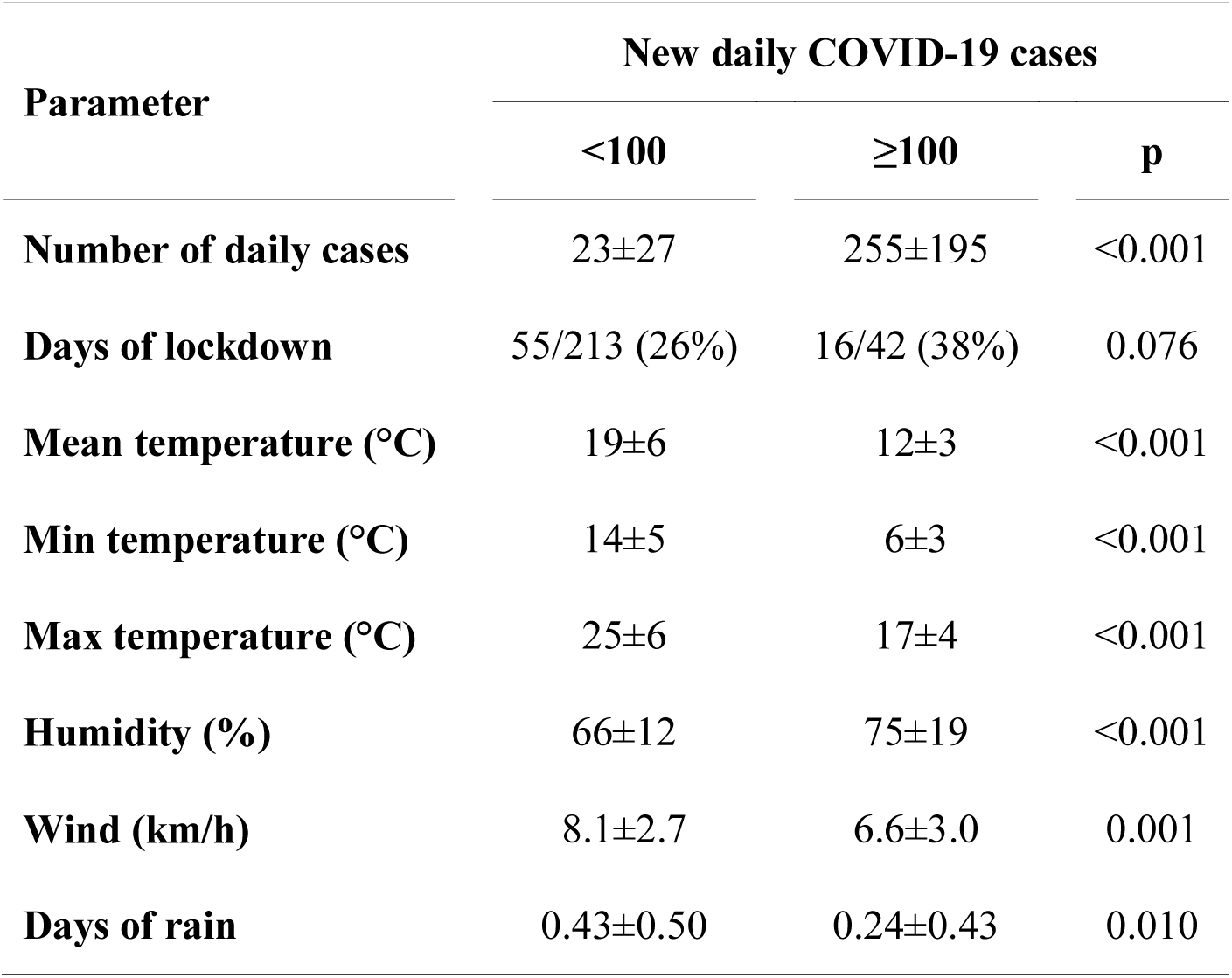
Values of weather parameters recorded in Verona (Italy) between days with <100 or ≥100 new daily diagnoses of coronavirus disease 2019 (COVID-19) in the same area, between March 1 and November 11, 2020.

## Discussion

Controversy remains as to whether some climate factors may significantly impact SARS-CoV-2 community transmission, especially in crowded settings and environments (12). To provide further insights on this critical issue, we carried out a retrospective study, arbitrarily limited to Verona (Italy), for increasing the accuracy of climate measures and the strength of their potential associations with COVID-19 epidemics. In this way, it is more likely that the weather parameters recorded in the area of Verona, which extends over 3,096 km^2^ and has a population of nearly 930,000 inhabitants, would help to more accurately discern the interplay with virus transmission compared to studies based on larger metropolitan areas, bigger districts, regions or even on entire nations or continents, where climate condition may be considerably different from one location and another.

Taken together, the results of our analysis reveal that the new daily COVID-19 diagnoses in Verona were positively associated with local humidity, while an inverse correlation was noted with mean, min and max temperature, mean wind speed and number of rainy days. Therefore, although each of these weather parameters that we have investigated seem to produce a direct effect on interpersonal SARS-CoV-2 spread, the cumulative number of daily contagions appears to be more dependent on mean air temperature, humidity, mean wind speed and rain, which altogether would contribute to explain over 60% of total variance in the number of new daily COVID-19 diagnoses in Verona (Table 2).

In a recent meta-regression of data garnered from 33 large US cities (i.e., with over 500,000 inhabitants), Takagi et al. concluded that the actual prevalence of COVID-19 was inversely associated with UV index, sun hours and outdoor temperature (13), whilst a positive association was found with mean wind speed. These findings are hence aligned with our results. In a separate investigation, Shi et al. explored the potential relationship between daily confirmed cases of COVID-19 and mean outdoor temperatures in 31 regions in mainland China (14). Overall, a significant inverse association could be found between air temperature and COVID-19 incidence (Coefficient = -0.01). Liu et al. also carried out a study of 30 provincial capital cities in China (15), concluding that meteorology may play an important role in the spread of COVID-19. More specifically, each 1 °C increase in outdoor temperature and diurnal temperature range was associated with 20% and 10% reduction in the number of daily diagnoses of COVID-19, respectively. Unlike these findings, a recent study published by Su et al. based on analysis of variation of effective reproductive number (Rt) among 277 worldwide regions, concluded that outdoor temperature is modestly associated with transmission rate, accounting for only 2.7% variation per 1 °C (16). Although this evidence persuaded the authors to conclude that warm temperatures are highly unlikely to prevent the natural spread of SARS-CoV-2 within the population, this conclusion, based only on temperature, seems inappropriate since warmer seasons are characterized by several climate changes (e.g., humidity, rainfall and so forth), which may all contribute to influence SARS-CoV-2 interpersonal spread.

The low risk of being infected by SARS-CoV-2 with increasing mean outdoor temperature values has at least three substantial explanations. First, temperate likely directly affects social distancing, since people are more inveigled to spend time outdoor, thus reducing the higher risk attributable to indoor viral transmission (17). Second, longer sunlight exposure and higher outdoor temperature have also been associated with greater extent of viral inactivation (18, 19). Finally, reliable evidence has been provided that virus-containing droplets could travel 3-fold slower in higher temperature environments (20), thus consistently mitigating their air propagation and infective potential.

Background airspeed seems to be another important determinant of infectious disease transmission. Although it is quite understandable that viral-containing aerosols and droplets may propagate over longer distances as the airflow speed increases, it has also been demonstrated that higher air velocity may effectively dilute the number of infectious viral-containing particles (7, 20). In keeping with this theory, a decreased wind speed has been inversely associated with the number of COVID-19 cases in five major Saudi Arabian cities (21). Since the risk of being infected and developing severe COVID-19 illness is directly dependent on the volume of viral inoculum (22), it is predictable that a lower risk of viral transmission would occur in environments with higher air currents and/or enhanced ventilation (23).

We also found an interesting association between days with rainfall and emergence of new COVID-19 cases in the area of Verona. The likelihood of diagnosing more SARS-CoV-2 cases seemed to be greater during days without precipitations. A similar association was reported in a recent study carried out by Chien and Chen in the top 50 US counties (24). These authors concluded that the risk of SARS-CoV-2 infection may considerably decline when the volume of yearly precipitation increases to over 450 cm. The mechanisms underneath this intriguing association between rainfall and lower risk of SARS-CoV-2 infection are still unclear at this point. We proffer that precipitations may reduce viral transmission by enhancing the fall of droplets and aerosol to the ground, thus limiting their airborne propagation. The drops of rain could also be effective in diluting, cleaning, or even completely washing away viable viral particles from infected outdoor surfaces, thus contributing to a lower risk of contagion contact.

Finally, concerning humidity, Zhao et al. recently showed that air transmission of virus-containing droplets is directly associated with humidity, increasing almost in parallel with the extent of environment humidity (20). Interestingly, these same authors concluded that the current 1 to 2 meters social distance standards might be insufficient in many ambient conditions, because droplets may travel far beyond such distances in extremely humid and cold environments.

## Conclusions

Although we could not directly evaluate the potential impact of the total number of daily molecular tests on the rate of new COVID-19 diagnoses in Verona, the evidence garnered from our analysis of meteorological data throughout the current SARS-CoV-2 local outbreak suggests that climate conditions may play an important role in modulating the conditions of viral transmission, thus influencing the likelihood or course of local infective clusters. Public health policies, preventive measures, testing strategies and hospital preparedness should be reinforced during periods of higher meteorological risk and in local environments with adverse climate conditions.

## Data Availability

Data will be available upon request

## Footnote

### Ethical Statement

The authors are accountable for all aspects of the work in ensuring that questions related to the accuracy or integrity of any part of the work are appropriately investigated and resolved. This retrospective analysis was based on unrestricted database searches, and thereby no patient’s informed consent or Ethical Committee approvals are necessary.

### Open Access Statement

This is an Open Access article distributed in accordance with the Creative Commons Attribution-NonCommercial-NoDerivs 4.0 International License (CC BY-NC-ND 4.0), which permits the noncommercial replication and distribution of the article with the strict proviso that no changes or edits are made and the original work is properly cited (including links to both the formal publication through the relevant DOI and the license). See: https://creativecommons.org/licenses/by-nc-nd/4.0/.

